# Deciphering the complex interplay between physical activity, inflammatory bowel disease and obesity/BMl through causal inference and mediation analyses

**DOI:** 10.1101/2024.04.15.24305823

**Authors:** Wensuo Lian, Yuhui Zhao, Hongxia Li, Linlin Jia, Kaixin Yao, Nan Li, Yuxin He

## Abstract

This study investigated the causal effect of physical activity (PA) on inflammatory bowel disease (IBD) and the role of body mass index (BMI) as a mediator. We used instrumental variables for moderate to vigorous physical activity (MVPA) during leisure time and leisure screen time (LST) derived from genome-wide association studies (GWASs) meta-analysis. IBD, Crohn’s disease (CD), and ulcerative colitis (UC) summary statistics were sourced from GWASs on European populations. We conducted univariable Mendelian randomization (UVMR) to assess the direct impacts of MVPA and LST on IBD risk. Multivariable Mendelian randomization (MVMR) was used to evaluate the mediating effects of BMI. Results indicated a protective effect of MVPA against IBD and CD. Conversely, higher genetically-predicted LST correlated with increased IBD and CD risks. BMI mediated 0.8% and 3.7% of the LST effect on IBD/CD, respectively, and 3.5% and 11.0% of the MVPA effect. Thus, PA appears causally related to lower IBD/CD risk, with BMI partially mediating this relationship, highlighting the risk increase from reduced PA via elevated BMI.

**Key messages:** *What is already known?:* Observational studies have shown that occupations involving more physical labor are associated with a lower risk of IBD compared with sedentary occupations.

*What is new here?:* The causal effect of PA on IBD and the role of BMI as a mediator between PA and IBD were investigated.

*How can this study help patient care?:* The incidence of CD can be reduced by promoting lifestyle management, such as reducing recreational sedentary activities and encouraging proper exercise. In addition, individuals who are not physically active should monitor their BMI to prevent the development of CD.

## 1. Introduction

Inflammatory bowel disease (IBD), including Crohn’s disease (CD) and ulcerative colitis (UC), are chronic and life-threatening inflammatory diseases^1^. CD manifests as transmural inflammation and longitudinal ulceration and affects the entire gastrointestinal tract, in particular the terminal ileum and adjacent colon, whereas UC manifests as mucosal inflammation and is restricted to the colon^2,3^. In recent years, breakthroughs in our knowledge of the pathogenesis of these diseases have resulted in the availability of a wider range of therapeutic options for IBD; however, owing to the complex pathogenesis and lack of knowledge regarding the exact cause of the diseases, there exists a therapeutic ceiling^4^. Currently, CD and UC are incurable, and patients often require lifelong treatment with drugs^5^. To improve the quality of life of patients with IBD, determining potential modifiable risk factors and treatments to slow down the progression or even prevent the onset of these diseases is important.

Recently, physical activity (PA) has attracted extensive attention as one of the most prevalent modifiable risk factors for diseases worldwide. PA is defined as any mechanical movement produced by skeletal muscular action that results in energy expenditure above basal levels^6^. Early studies have shown that occupations involving more physical labor are associated with a lower risk of IBD compared with sedentary occupations^7,8^. In two large prospective cohorts of American women, higher levels of PA were associated with a reduced risk of CD compared to lower levels of PA, but no association was found between UC and PA^9^. A meta-analysis also came to the same conclusion^10^; however, a study by Chan et al. reported a different result, i.e.^11^, PA did not show any association with UC or CD. These contradictions in the observational studies indicate that additional studies are required to estimate the actual causal relationships. Furthermore, many observational studies have reported that physical inactivity and increased sedentary time are associated with an increased risk of obesity^12,13^. Meanwhile, a substantial and consistent body of epidemiological data suggests that obesity is one of the high-risk factors for developing IBD^14,15^. These findings indicate that the increased risk of CD observed in individuals who engage in lower levels of PA may be partially mediated by weight gain. However, to date, no study has validated this hypothesis.

Traditional observational studies form the backbone of our current understanding of the mediating pathways; however, these studies might have been influenced by potential confounding, measurement error, and reverse causality. Thus, the extent to which the association observed between exposure and outcome and its intermediates is influenced by bias, such as confounding, measurement error, and reverse causality, remains unclear. These limitations can be overcome by applying Mendelian randomization (MR), which is an accepted method of investigating causal relationships in observational data. MR uses single-nucleotide polymorphisms (SNPs) elucidated by genome-wide association studies (GWASs) to be strongly associated with an exposure as instrumental variables (IVs) to determine whether a causal relationship exists between an exposure and outcome. Since genetic variants are randomly allocated at conception prior to disease onset, MR analysis efficiently avoids potential confounding factors and reverse causality^16^. As a recent development in the MR, multivariate MR (MVMR) method is used to investigate mediation.

In this study, we aimed to determine potential causal relationships between self-reported moderate to vigorous intense PA during leisure time (MVPA)/leisure screen time (LST) and the risk of IBD, including that of its two subtypes, i.e., CD and UC, by analyzing the most recent GWAS data using the univariable two-sample MR (UVMR) method. Furthermore, using the MVMR method, we aimed to determine whether BMI mediates the effects of MVPA/LST on IBD, including CD and UC, and the proportion of the effect that is mediated by BMI.

## 2. Methods

### 2.1 Data Sources

#### 2.1.1 Summary-level data of MVPA and LST

Summary statistics of the main exposures analyzed in this study, i.e., MVPA (Moderate to vigorous intense physical activity during leisure time) and LST (leisure screen time), were extracted from the latest published meta-analysis of GWASs on self-reported MVPA and LST. The meta-analysis analyzed 51 GWASs and included a total of 703,901 European individuals (MVPA: n = 608,595; LST: n = 526,725)^17^ (summarized in Table 1). Activities such as swimming and jogging were regarded as MVPA, and activities such as watching TV, playing video games, and sitting at the computer were regarded as LST. The results of the GWASs were adjusted for principal covariates, including assessment center, genotyping array, age, and season.

**Table 1:** Characteristics of GWAS included in MR analyses.

| Trait/phenotype | GWAS consortium | Ethnicity | Sample size (total or case/control subjects) | PMID |
| --- | --- | --- | --- | --- |
| MVPA | MTAG | European | 608,595 | 36071172 |
| LST | MTAG | European | 526,725 | 36071172 |
| IBD | IBD meta-GWAS | European | 25,042/34,915 | 28067908 |
| CD | CD meta-GWAS | European | 12,194/28,072 | 28067908 |
| UC | UC meta-GWAS | European | 12,366/33,609 | 28067908 |
| BMI | GIANT | European | 681,275 | 30124842 |

GWAS data sets included in MR analyses. PMID, PubMed identifier. Abbreviations: MVPA, moderate to vigorous intense physical activity during leisure time. LST, leisure screen time. IBD, inflammatory bowel disease. CD, Crohn's disease. UC, ulcerative colitis. BMI, Body Mass Index.

#### 2.1.2 Summary-level data of IBD

Summary statistics of IBD were extracted from a meta-analysis of GWASs of IBD that included a total of 25,042 clinically diagnosed cases of IBD (with 34,915 controls), 12,194 clinically diagnosed cases of CD (with 28,072 controls), and 12,366 clinically diagnosed cases of UC (with 33,609 controls) (summarized in Table 1). The first 10 principal components for each cohort were adjusted^18^.

#### 2.1.3 Summary-level data of BMI

Genome-wide significant (P < 5 × 10^-8^) genetic variants associated with BMI were extracted from a meta-analysis of GWASs, which included a total of 681,275 individuals of European ancestry, published by the Genetic Investigation of Anthropometric Traits consortium (summarized in Table 1). Age, age squared, and study-specific covariates, including principal components for adjusting for population stratification, were adjusted^19^.

### 2.2 Selection of IVs

For MR analysis, we included IVs that met the following three assumptions (Fig 1): (a) the IV must be robustly associated with the exposure; (b) the IV must be unrelated to any known confounders; and (c) the IV must be associated with the risk of outcome only via its effects on the exposure. First, SNPs associated with PA were extracted, with the genome-wide significance threshold as P < 5 × 10^-8^ for MVPA and LST. Second, SNPs that satisfied the relevance assumption (r^2^ = 0.01, clumping distance = 10,000 kb) were regarded as being correlated with the MVPA and LST and to exhibit genome-wide significance. Additionally, they were validated to be independent and not in linkage disequilibrium. Moreover, only data pertaining to individuals of European ancestry were analyzed in this study, and population stratification did not violate the independence assumption at any instance.

**Fig 1:**
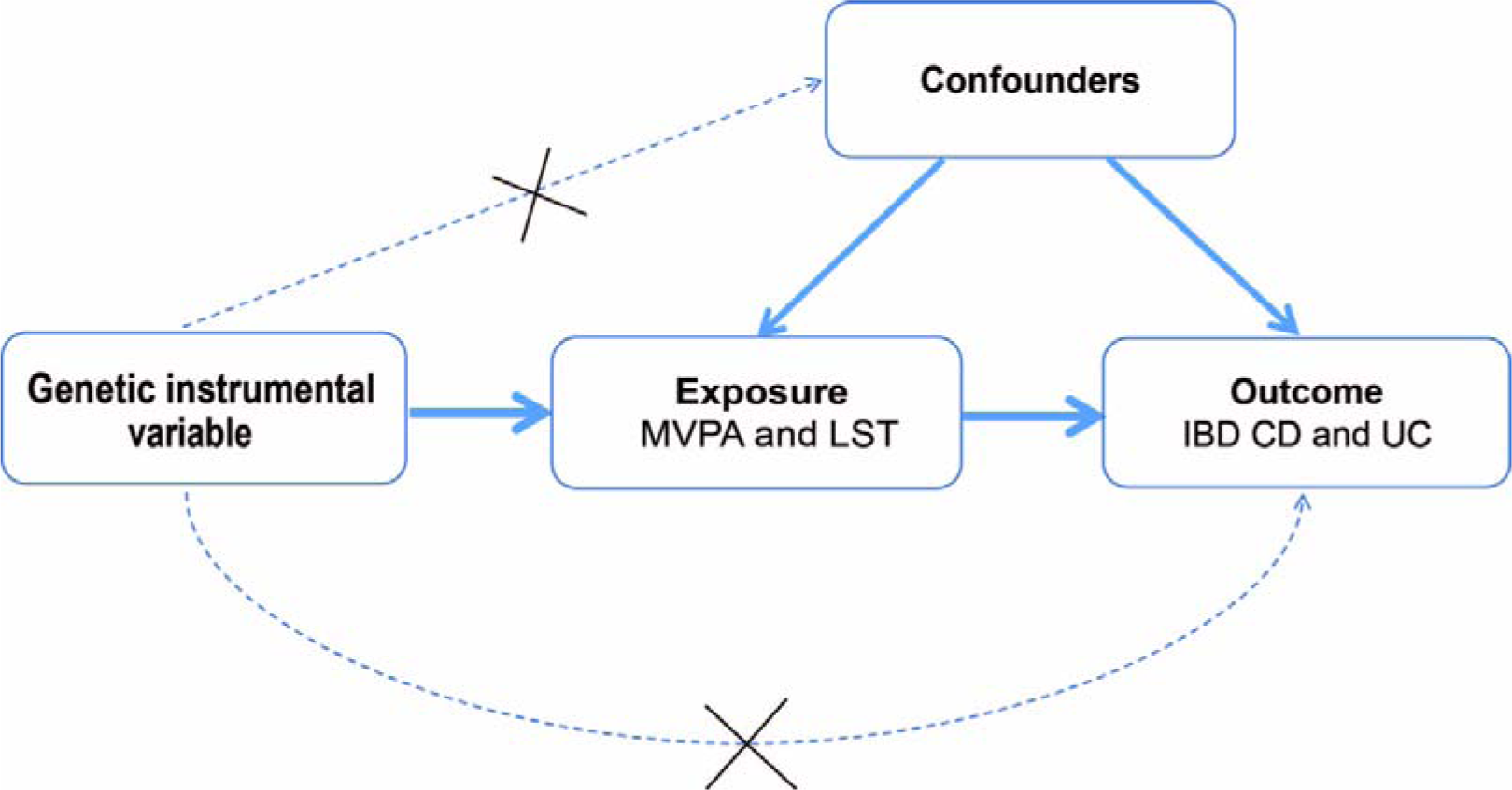
Mendelian randomization model of exposure and outcome. The design is under the assumption that the genetic instrumental variables are associated with exposure, but not with confounders, and the genetic instrumental variable influence outcome only through exposure. Genetic instrumental variable indicates single nucleotide polymorphism.

### 2.3 Statistical analysis

#### 2.3.1 Total, direct, and indirect effects

First, UVMR analysis was performed to determine the net effect of MVPA and LST on IBD (including CD and UC), which was regarded as the total effect. Next, UVMR analysis was performed again to estimate the effect of MVPA and LST on BMI. Subsequently, MVMR analysis was performed to determine the effect of mediators on IBD (including CD and UC) after adjusting for MVPA and LST.

The total effect of an exposure on an outcome can be broken down into direct and indirect effects. Direct effect is defined as the effect of exposures on an outcome that is not mediated through a causal intermediate, whereas indirect effect is defined as the effect of an exposure on an outcome through a candidate mediator (Fig 2). Herein, the indirect effect was calculated using the product of the coefficients method, wherein, after adjusting for MVPA and LST, the MR estimate of the effect of MVPA and LST on BMI was multiplied with the MR estimate of the impact of BMI on IBD (including CD and UC). Furthermore, the proportion of the effect mediated by BMI was calculated by dividing the indirect effect by the total effect. The confidence intervals (CIs) and standard errors for the proportion of the effect mediated by BMI were estimated using the delta method.

**Fig 2:**
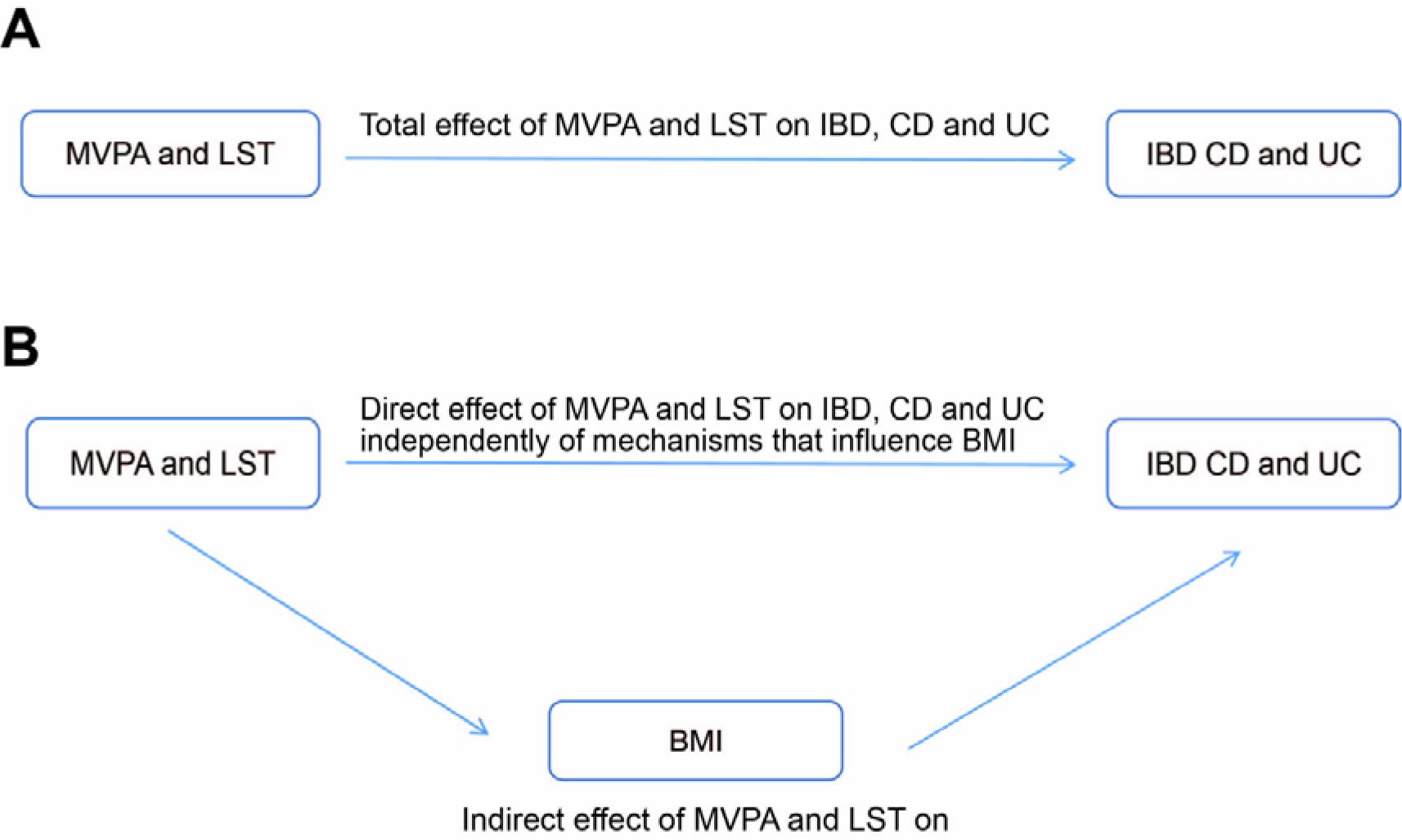
Direct acyclic graph to illustrate total, direct, and indirect effects of MVPA and LST on IBD, CD, and UC. Directed acyclic graphs demonstrating the hypothesized direction for the total effect of MVPA and LST on IBD, CD and UC (A) and the hypothesized direction for the effect of MVPA and LST on BMI (B), which may partially mediate the effect of MVPA and LST on IBD, CD and UC.

#### 2.3.2 UVMR and MVMR analysis

To evaluate the effect of MVPA and LST on the risk of IBD (including that of CD and UC), we used the fixed-effects inverse variance-weighted (IVW) method as the main UVMR and MVMR method. The IVW method is used to estimate the odds ratio (OR) for genetic variants and is the most widely used method in MR studies. Herein, for IVW analysis, we used the Wald estimator method to estimate the causal effect of each SNP and determined the overall effect by meta-analyzing all the estimates.

#### 2.3.3 MR sensitivity analyses

Sensitivity analyses were performed to validate the robustness of the IVW results and determine pleiotropy and the potential genetic outliers. A weighted median estimator method, which is a modification of the standard weighted median MR, was employed to examine the median effects of all the SNPs and resulting in unbiased estimates of effects under the assumption that >50% of the information contributing to the analysis comes from valid IVs. MR–Egger method was used to test directional pleiotropy and causal effects that on average differed from zero and to estimate the causal effects under the InSIDE (Instrument Strength Independent of Direct Effect) assumption. In MR–Egger regression, the slope is used to estimate the causal effect of the exposure on the outcome when it is consistent with the strength of the instrument independent of the direct effects assumption; in addition, deviation of the intercept of MR–Egger regression from zero indicates the presence of pleiotropy. Despite the statistically lower efficiency (i.e., a wider range of CIs), the MR–Egger method provides a causal estimate (i.e., regression slope) corrected for directional horizontal pleiotropy. The MR pleiotropy residual sum and outlier method (MR PRESSO) was employed to detect significant outliers, and outliers were removed to correct horizontal pleiotropic effects. The global test was used to assess whether horizontal pleiotropy existed among the instruments. Furthermore, Cochran Q test was used to evaluate the heterogeneity among the causal effects of each variant. P < 0.10 was regarded to indicate statistically significant heterogeneity. We also performed “leave-one-out” analysis to determine and assess the potential impact of specific variants on the estimates by excluding one SNP each turn and performing IVW analysis on the other SNPs.

The results of the MR analyses are represented as ORs, and 95% CIs were determined for the risk of outcomes for the corresponding unit changes in exposure. The OR of the outcome risk per log OR change in exposure was interpreted as the final effect estimates, given that all exposure variables in the current analysis were binary. MR analyses were performed using the ‘‘TwoSampleMR” package in the R software (version 4.1.2).

## 3. Results

### 3.1 Total and direct effects of MVPA and LST on IBD, CD, and UC

In UVMR, IVW analysis suggested a protective causal relationship between MVPA and IBD as well as MVPA and CD (Table 2, Fig 3 A-B). For each S.D. increase in MVPA, the relative odds of IBD decreased by 33.5% (OR = 0.665, 95% CI = 0.465–0.952, P = 0.026) and the relative odds of CD decreased by 46.7% (OR = 0.533, 95% CI = 0.302–0.942, P = 0.030). IVW analysis showed that a higher genetically-predicted LST was associated with increased risk of IBD and CD (Table3, Supplementary Fig 1 A-B). For each S.D. increase in LST, the relative odds of IBD (OR = 1.213, 95% CI = 1.063–1.384, P = 0.004) increased by 21.3% and the relative odds of CD (OR = 1.245, 95% CI = 1.070–1.449, P = 0.005) increased by 24.5%. On the contrary, MVPA did not show a statistically significant association with a reduced risk of developing UC (OR = 0.761, 95% CI = 0.489–1.183, P = 0.225) (Table 2, Fig 3 C). Furthermore, no statistically significant evidence of a relationship was observed between LST and UC (OR = 1.122, 95% CI = 0.950–1.325, P = 0.176) (Table 3, Supplementary Fig 1 C). Similar causal estimates were observed for IBD (including CD and UC) using the other MR methods, including the weighted median and MR–Egger methods.

**Fig 3:**
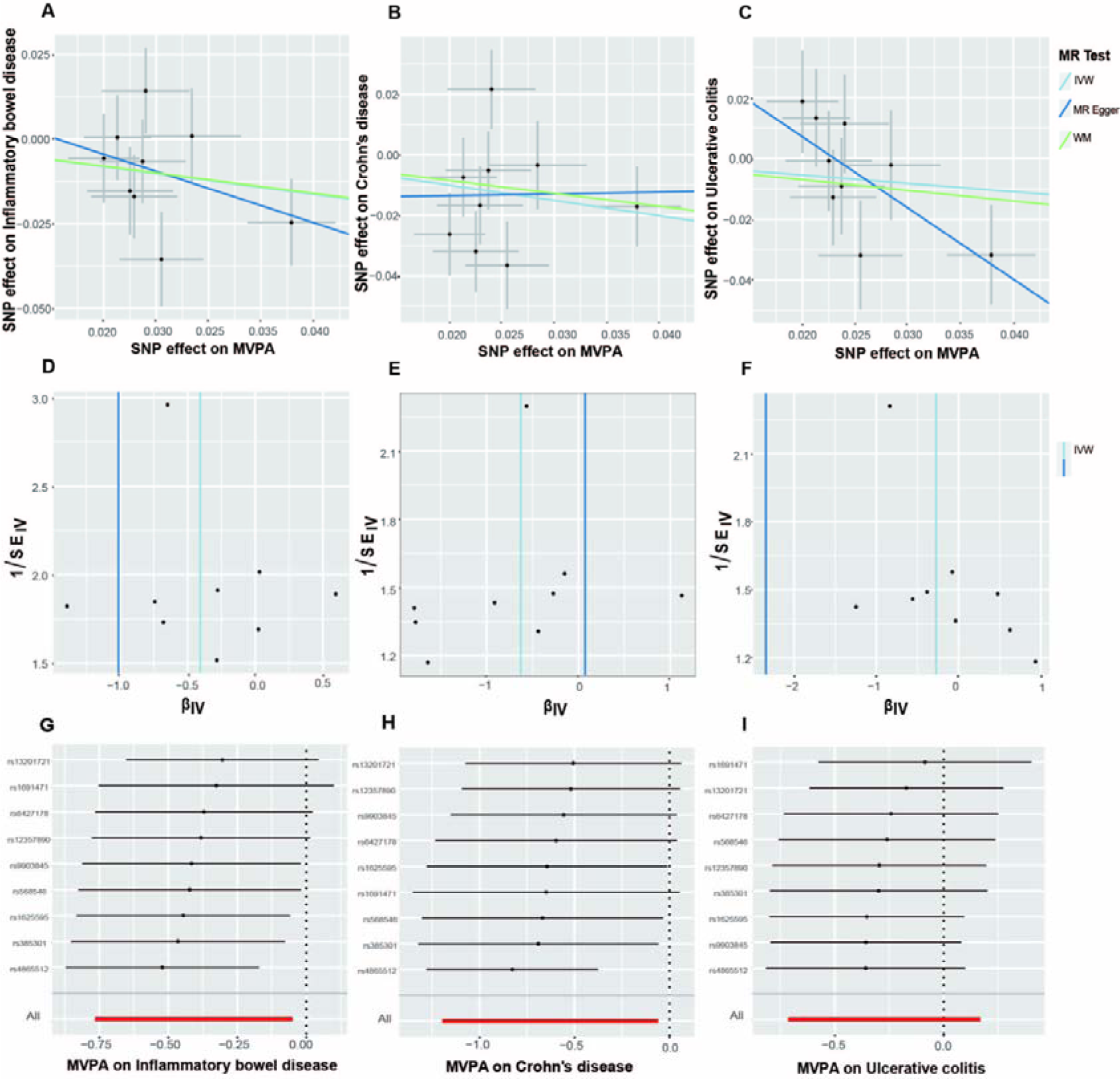
Scatter plots, funnel plots and leave-one-out analysis for the association of MVPA on IBD, CD and UC. (A-C) Scatter plots for IVW, MR-Egger and WM analysis methods demonstrating the effect of MVPA on IBD (A), CD (B) and UC (C). (D-F) Funnel plot of MVPA on IBD (D), CD (E) and UC (F) to suggest no evidence of substantial heterogeneity. (G-I) Leave-one-out analysis to explore whether the causal link of MVPA on IBD (G), CD (H) and UC (I) was driven by single specific SNP.

**Table 2:.** The causal effect of MVPA on IBD, CD, and UC.

| exposur<br>e | outcome | SNPs | MR methods | OR | 95%CI | Pval | Q_pval | Intercept_pval |
| --- | --- | --- | --- | --- | --- | --- | --- | --- |
| MVPA | IBD | 9 | IVW | 0.665 | 0.465,0.952 | 0.026 | 0.315 | 0.538 |
|  |  |  | MR-Egger | 0.364 | 0.056,2.353 | 0.324 | 0.267 |  |
|  |  |  | WM | 0.671 | 0.424,1.062 | 0.088 |  |  |
|  | CD | 9 | IVW | 0.533 | 0.302,0.942 | 0.030 | 0.079 | 0.654 |
|  |  |  | MR-Egger | 1.076 | 0.054,21.636 | 0.963 | 0.057 |  |
|  |  |  | WM | 0.588 | 0.320,1.083 | 0.089 |  |  |
|  | UC | 9 | IVW | 0.761 | 0.489,1.183 | 0.225 | 0.368 | 0.093 |
|  |  |  | MR-Egger | 0.096 | 0.011,0.809 | 0.068 | 0.668 |  |
|  |  |  | WM | 0.704 | 0.396,1.254 | 0.234 |  |  |

The Q\_pval was executed by the MR-Egger and IVW methods to detect heterogeneity. The intercept p-value was derived from the pleiotropy test of MR- Egger regression. Abbreviations: MVPA, moderate to vigorous intense physical activity during leisure time. IBD, inflammatory bowel disease. CD, Crohn's disease. UC, ulcerative colitis.

**Table 3:.**
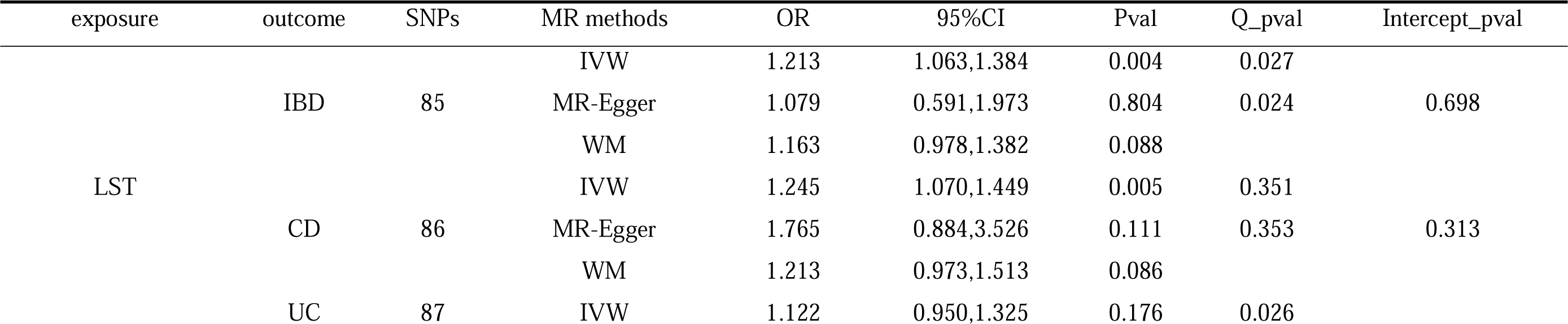
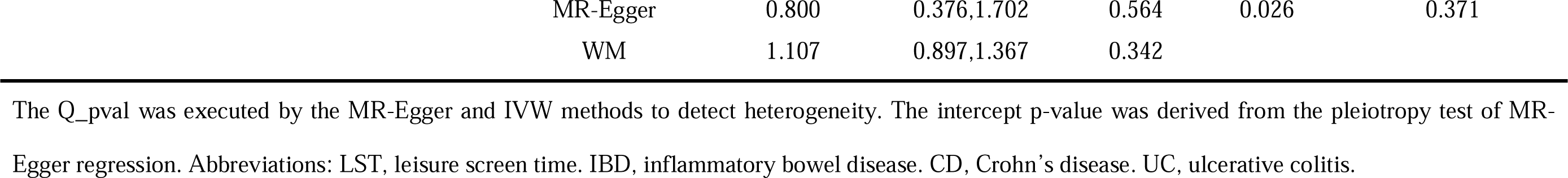
The causal effect of LST on IBD, CD, and UC.

In the MVMR analyses, a causal relationship between MVPA and IBD was observed after adjusting for BMI. For each S.D. increase in MVPA, the relative odds of IBD decreased by 32.3% (OR = 0.677, 95% CI = 0.464–0.986, P = 0.042) and the relative odds of CD decreased by 42.2% (OR = 0.578, 95% CI = 0.356–0.936, P = 0.026) (Table 4). On the contrary, the causal relationship between LST and IBD (OR =1.074, 95% CI = 0.831–1.388, P = 0.586) or LST and CD (OR = 1.107, 95% CI = 0.797–1.537, P = 0.543) did not show statistical significance after adjusting for BMI (Table 4).

**Table 4:**
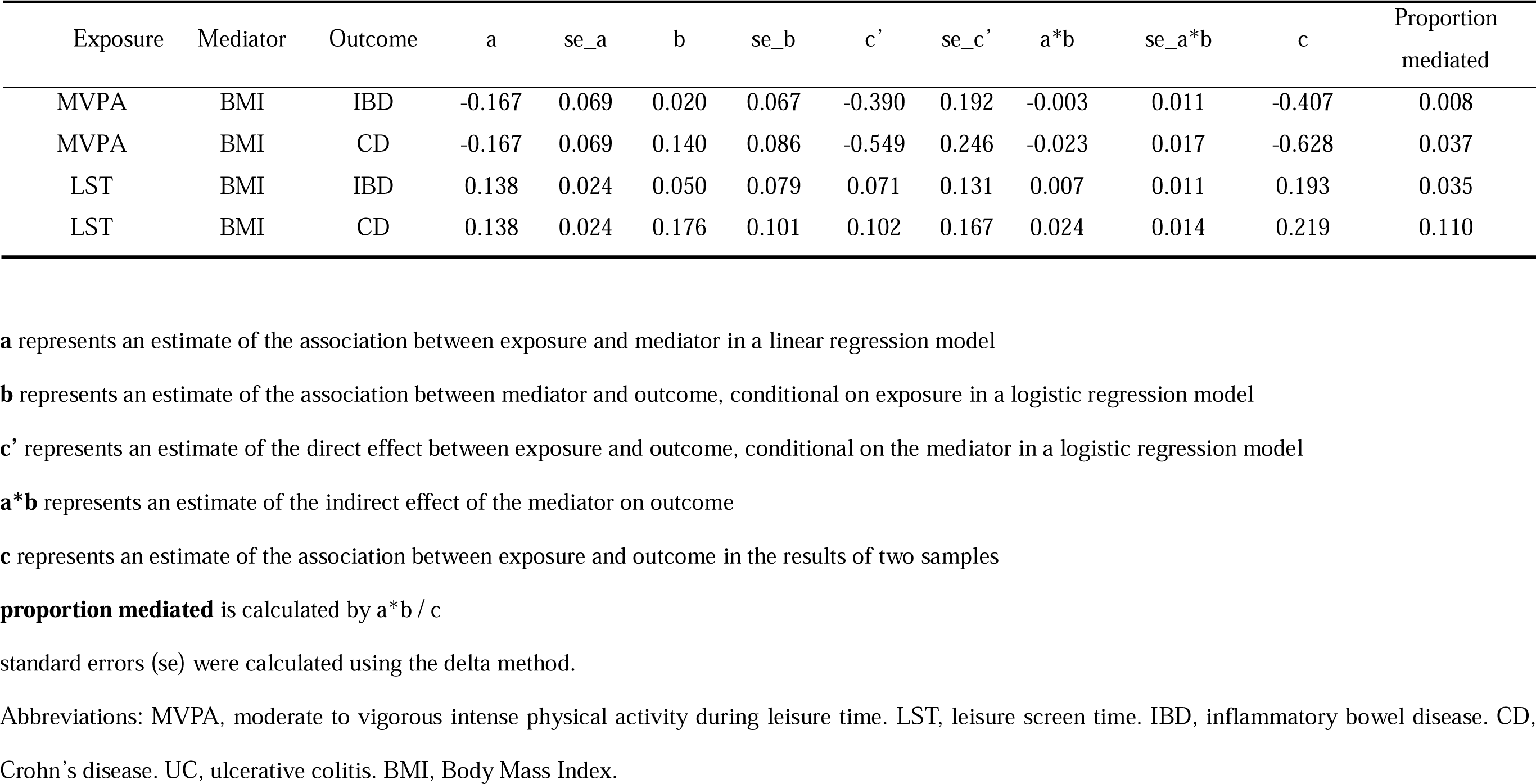
Mediation analysis in our research.

### 3.2 Effect of MVPA and LST on BMI

To explore the effect of MVPA and LST on IBD (including CD and UC) mediated through BMI, we performed UVMR analysis to estimate the effect of MVPA and LST on BMI. The results showed that higher levels of MVPA were associated with lower BMI and that higher levels of LST were associated with higher BMI. For each SD increase in MVPA, the BMI was observed to decrease by 0.154 SD units (OR = 0.846, 95% CI = 0.739–0.969, P = 0.016), and for each SD increase in LST, the BMI was observed to increase by 0.148 SD units (OR = 1.148, 95% CI = 1.094–1.204, P = 1.48×10^-8^).

### 3.3 Effect of BMI on IBD, CD, and UC after adjusting for MVPA and LST

In the MVMR analyses of MVPA–BMI–IBD and MVPA–BMI–CD, the conditional F-statistics for MVPA and BMI were both >10. After accounting for MVPA, the direct effect of BMI on IBD was determined to be OR 1.020 (95% CI = 0.894–1.163, P = 0.769), and the proportion mediated by BMI was determined to be 0.8%. Likewise, after accounting for MVPA, the direct effect of BMI on CD was determined to be OR 1.150 (95% CI = 0.971– 1.361, P = 0.104), and the proportion mediated by BMI was determined to be 3.7%.

In the MVMR analyses of LST–BMI–IBD and LST–BMI–CD, the conditional F-statistics for LST and BMI were both >10. After accounting for LST, the direct effect of BMI on IBD was determined to be OR 1.051 (95% CI = 0.901–1.226, P = 0.528), and the proportion mediated by BMI was determined to be 3.5%. After accounting for LST, the direct effect of BMI on CD was determined to be OR 1.192 (95% CI = 0.979–1.452, P = 0.081), and the proportion mediated by BMI was determined to be 11.0% of the total effect (Table 4).

### 3.4 Sensitivity analyses

We performed a series of sensitivity analyses, including the MR–Egger intercept test, Cochran Q test, and MR–PRESSO global test, to assess the robustness of our results. In the MR–Egger intercept tests, P values > 0.05 were not likely to exhibit horizontal pleiotropy. Even though heterogeneity was observed in the Cochran Q test, the MR estimates did not invalidate random-effect IVW. Furthermore, the pooled heterogeneity was balanced. Altogether, our results were less likely to be confused by horizontal pleiotropy, heterogeneity (Fig 3 D-F, Supplementary Fig 1 D-F), or individual SNP effects, as demonstrated by the leave-one-out analyses (Fig 3 G-I, Supplementary Fig 2 A-C).

## 4. Discussion

In this study, we used the UVMR and MVMR methods to analyze data from large-scale GWASs. First, our results suggest an effect of genetically-predicted reduced MVPA, increased LST, and increased BMI on the risk of IBD and CD. However, our results suggest no association between PA/ LST and risk of UC. Second, our study provides robust evidence that the effect of MVPA/LST on IBD and CD is partially mediated by BMI. Altogether, our study reveals the complex association between MVPA/LST, BMI, and IBD, including its subtypes; moreover, our findings address the gap in knowledge on this topic in the general population.

Several observational studies have investigated the association between PA and the risk of IBD. The Nurses’ Health Study I and II reported that women aged between 25 to 55 years who engaged in at least 27 metabolic equivalent tasks (MET) hours of PA per week exhibited an HR of 0.56 (0.37; 0.84) for the risk of developing CD compared with women who were inactive; however, these studies reported no association between PA and the risk of UC^9^. These findings are consistent with those of our study. However, a study on the European Prospective Investigation into Cancer and Nutrition cohort did not report an association between PA and the risk of CD and UC^11^. These conflicting results may be because the cohort included only 75 CD cases and 177 UC cases and because the study was unable to account for long-term changes in PA. Moreover, this contradictory result may be attributed to the analysis method itself. Observational studies may be influenced by unavoidable clinical confounders, which influence the exposures and outcomes, thereby making it difficult to accurately determine causal relationships.

Despite long-standing clinical observations^11,20^, the causal genetic association between BMI and CD was not discovered until recently^21^. However, no previous study has investigated whether increased BMI mediates the effect of MVPA/LST on the risk of CD. The results of our study suggest an indirect effect of MVPA/LST on the risk of CD that is mediated by BMI. Furthermore, in the context of MR, this effect is highly unlikely to have been confounded by other factors. Our MR analysis indicates that approximately 4% of the effect of reduced MVPA on increased CD risk is mediated by BMI and that approximately 11% of the effect of increased LST on increased CD risk is mediated by BMI. The difference in the mediated proportion may be attributed to the fact that compared to reduced MVPA, increased LST results in lower energy expenditure and higher energy intake (particularly snacking), thereby resulting in a greater shift in energy balance to excess energy and a more significant increase in BMI^22,23^.

Our in-depth analyses of the mediating effect of BMI elicited potential explanations for this causal association between MVPA/LST and IBD/CD. Inflammatory T cells are known to direct innate cells to maintain a constant hypersensitivity to microbial antigens, tissue injury, and chronic intestinal inflammation. Therefore, the accumulation of inflammatory T cells in the intestine is considered to be one of the main pathogenic mechanisms of IBD^24^. A substantial body of research suggests that obesity may contribute to IBD and CD by altering innate and intrinsic immune responses. In obese individuals, adipocytes can synthesize and secrete several biologically active substances called adipokines, such as lipocalin adiponectin (APN), IL-1, IL-6, IL-8, IFNγ, TNF-α, and leptin^25,26^. Among these, leptin and lipocalin adiponectin are of particular interest. The balance between leptin and adiponectin, i.e., the leptin–adiponectin ratio, is important for the maintenance of intestinal immune homeostasis^26^. Leptin exerts a notable pro-inflammatory effect on the immune system and can be released when stimulated by inflammation^27^. In contrast, lipocalin has an inhibitory effect on the expression of adhesion molecules and pro-inflammatory mediators^26^. At the onset of obesity, the leptin–adiponectin ratio is altered, wherein the level of leptin is elevated. Increased leptin secretion induces the proliferation of inflammatory CD4 T cells and results in an increase of type 1 T helper (Th1) cells and the suppression of Th2 cytokines^28^. In addition, TNF-α, IL-1β, and IL-6 have been demonstrated to recruit CD4 T cells and enhance Th1/17 immunity; moreover, IFN-γ has been shown to induce Th17 cell development^29-31^.

IBD may be caused by inflammatory T cells influencing the function of innate cells, such as epithelial cells, fibroblasts, and phagocytes, which results in constant hyperresponsiveness to microbial antigens being activated, thereby causing intestinal tissue injury and chronic intestinal inflammation^24,32,33^. Under these conditions, intestinal permeability increases, and microbial components cross the barrier, inducing activation of DCs and macrophages, which in turn induce the infiltration of more inflammatory CD4 T-cells into the intestinal tissues^24,34^. Meanwhile, lipopolysaccharide present on the surface of microbes activates the NF-κB pathway, which results in the elevation of TLR-4 expression in adipocytes and preadipocytes, leading to increased production of adipokines, and creating a positive inflammatory feedback loop that further exacerbates the development of IBD^26,35^. Th1 and Th17-dominant immune cells have been shown to play a central role in the pathogenesis of CD, whereas Th2-type immune cells were shown to play an important role in UC^36^. This may explain why the occurrence of obesity leads mainly to IBD and CD rather than UC.

Taken together, our findings validated that increasing moderate to vigorous intense PA and reducing LST prevent CD and that this effect is partially mediated by BMI. Therefore, the incidence of CD can be reduced by promoting lifestyle management, such as reducing recreational sedentary activities and encouraging proper exercise. In addition, individuals who are not physically active should monitor their BMI to prevent the development of CD. This practice can reduce the risk of CD development as well as the burden on healthcare and general public health returns in terms of human productivity^37^.

This study has several strengths. First, this study is based on the largest and most up-to-date GWAS database on PA and is the first to use MR to analyze whether BMI mediates the effects of MVPA/LST on the risk of IBD. Second, we assessed the causal effects of MVPA and LST on the risk of IBD as a whole, as well as the two main subtypes of IBD, i.e., UC and CD, using an MR approach, which reduces unobserved confounding and is less susceptible to reverse causation and exposures that are non-differentially measured with error. Nevertheless, our study also has several limitations. First, the genetic instruments and GWASs used in this study were based on individuals of European ancestry; therefore, our results cannot be generalized to other ethnic groups. Second, our MR analysis only yielded genetic evidence. In the future, animal studies or population-based observational studies should be conducted to validate the causality, explore potential mechanisms of this possible causal association, and further explore related mechanisms. Third, the self-reported PA data may be affected by the participants’ memorizing and cognitive abilities, such as awareness of the beneficial effects of PA, which could have resulted in misclassification and outcome bias.

In conclusion, by leveraging large-scale genetic summary-level data, we found that elevated BMI partially mediates the effect of MVPA/LST on the risk of IBD and CD. Our study findings may provide a strategy for preventing IBD and CD. Further studies are warranted to decipher other potential mechanisms linking MVPA/LST to IBD and CD.

## Declarations

### Ethics approval and consent to participate

Since the analysis used published studies or publicly available GWAS summary data, containing no personal identifications, no ethical committee approval was required. The informed consent from participants, and approval by ethical committees involved in the original studies.

### Patient or volunteer consent

Not applicable.

### Consent for publication

Not applicable.

### Availability of data and materials

LST and MVPA data underlying this article are available in the NHGRI-EBI GWAS Catalog, and can be accessed with IDs GCST90104339 and GCST90104341, respectively. IBD, CD, UC and BMI data are available in the IEU OpenGWAS, and can be accessed with IDs ebi-a-GCST004131, ebi-a-GCST004132, ebi-a-GCST004133, ebi-b-40, respectively.

### Authors’ contributions

W.L., Y.Z., and Y.H. designed the study. W.L., Y.Z., H.L. and L.J. wrote the manuscript. W.L., Y.Z., K.Y. and N.L. acquired the data, assessed quality, analyzed and interpreted the data. All authors were involved in drafting of the manuscript and critical revision of the manuscript, including the authorship list. All authors approved the final draft of the manuscript. Prof. He had full access to all of the data in the study and takes responsibility for the integrity and the accuracy of the data analysis.

### Research registration unique identifying number

Not applicable.

## Data Availability

All data produced in the present work are contained in the manuscript.

## Acknowledgments

We are indebted to the staff and participants of the Genetic Investigation of Anthropometric Traits consortium, IEU OpenGWAS, the NHGRI-EBI GWAS Catalog and the genome-wide association study consortia for their important contributions, without whom this effort would not have been possible.

## Abbreviations

Inflammatory bowel disease (IBD); Crohn’s disease (CD); ulcerative colitis (UC); physical activity (PA); Mendelian randomization (MR); single-nucleotide polymorphisms (SNPs); genome-wide association studies (GWASs); instrumental variables (IVs); multivariate MR (MVMR); moderate to vigorous intense PA during leisure time (MVPA); leisure screen time (LST); univariable MR (UVMR); confidence intervals (CIs); inverse variance-weighted (IVW); odds ratio (OR); InSIDE (Instrument Strength Independent of Direct Effect); MR pleiotropy residual sum and outlier method (MR PRESSO); metabolic equivalent tasks (MET); type 1 T helper (Th1)

## Writing Assistance

Not applicable

## Synopsis

The main point of this article is that there is a causal relationship between PA and IBD/CD and that the portion of the increased risk of IBD/CD due to decreased PA is mediated by increased BMI.

