## Supplementary material for "Deciphering the complex interplay between physical activity, inflammatory bowel disease and obesity/BMl through causal inference and mediation analyses"

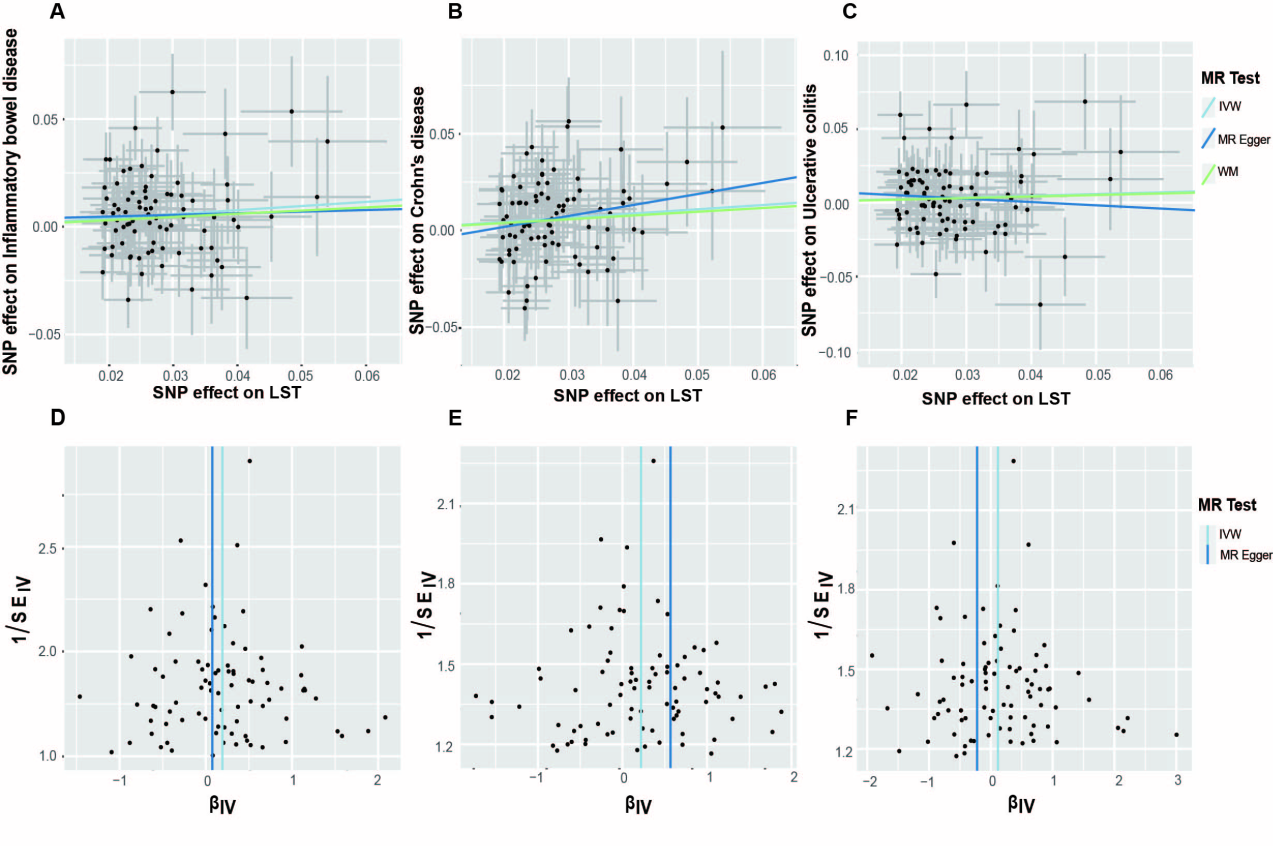
**Supplementary Fig 1: Scatter plots and funnel plots for the association of LST on IBD, CD and UC.** (A-C) Scatter plots for IVW, MR-Egger and WM analysis methods demonstrating the effect of LST on IBD (A), CD (B) and UC (C). (D-F) Funnel plot of LST on IBD (D), CD (E) and UC (F) to suggest no evidence of substantial heterogeneity.

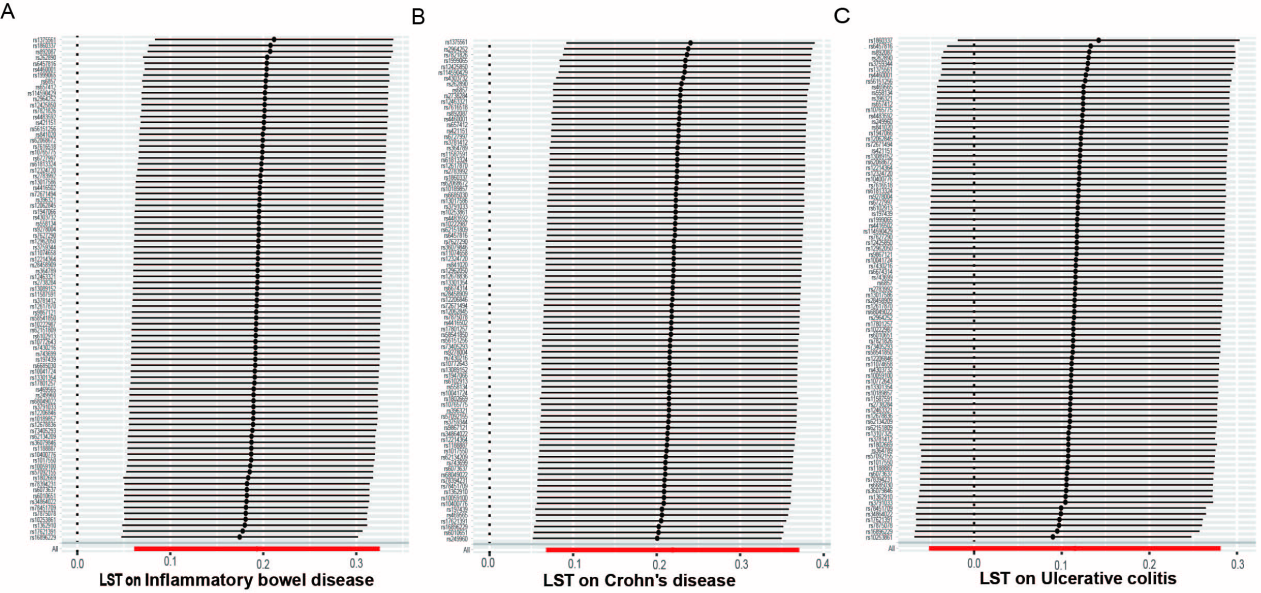

**Supplementary Fig 2: leave-one-out analysis for the association of LST on IBD, CD, and UC.** (A-C) Leave-one-out analysis to explore whether the causal link of LST on IBD (A), CD (B), and UC (C) were driven by a single specific SNP.
